# Novel mitochondrial variants associated with Parkinson’s disease reveal a high heteroplasmy-weighted polygenic risk score in Brazilians

**DOI:** 10.64898/2026.01.15.26344224

**Authors:** Gustavo Barra-Matos, Matheus Epifane-de-Assunção, Marcella Vitória Belém-Souza, Felipe Gouvea de Souza, Caio Santos Silva, Letícia Cota Cavaleiro de Macêdo, Tatiane Piedade de Souza, Giovanna C. Cavalcante, André Vitor de Souza Fernandes, Ândrea Ribeiro-dos-Santos, Bruno Lopes Santos-Lobato, Gilderlanio Santana-de-Araújo

## Abstract

Mitochondrial single-nucleotide variants (mtSNVs) can dysregulate cellular bioenergetics and have been increasingly implicated in susceptibility to Parkinson’s disease (PD). These variants may impair oxidative metabolism and respiratory chain efficiency, thus contributing to neuronal dysfunction and degeneration. In peripheral blood, mtSNVs may also reflect systemic immunometabolic alterations associated with PD; however, this aspect remains poorly explored, particularly in admixed populations with significant indigenous ancestry. In this study, we analyzed the complete mtDNA of peripheral blood samples from 179 admixed individuals (104 with PD and 75 controls) from the Brazilian Amazon. Associations between mtSNVs and PD were assessed using adjusted logistic regression models, and functional annotation was performed using the Variant Effect Predictor. Furthermore, we proposed and calculated a heteroplasmy-weighted mitochondrial polygenic risk score (mtPRS). We observed a higher mtSNV burden in PD patients, predominantly in *RNR2* gene and genes of Complexes I and IV. In total, 536 unique mitochondrial SNVs were identified (214 exclusive to PD, 321 shared between PD and controls, and one exclusive to controls). Four mitochondrial SNVs were associated with PD, including three novel variants (*COX1*: m.6630G>A, *COX2*: m.7613C>T, and *RNR2*: m.1996C>T) and one previously reported variant (*ND4*: m.12112C>T). Notably, the mitochondrial polygenic risk score (mtPRS) showed a strong association with increased PD risk (OR = 3.64; FDR = 1.12 × 10^-^_). Taken together, these results suggest that mtSNVs may contribute to PD susceptibility in admixed Amazonian population, highlighting the relevance of mitochondrial genetic architecture in PD and emphasizing the importance of including underrepresented populations in genomic research.

## 1 Introduction

Parkinson’s disease (PD) is the second most prevalent neurodegenerative disease in the world, characterized by the progressive degeneration of dopaminergic neurons in the *substantia nigra pars compacta* (SNpc) and by motor symptoms, as well as non-motor manifestations (Bloem et al. 2021). Although the pathophysiology of PD is multifactorial, involving both environmental and genetic factors, growing evidence suggests that alterations in mitochondrial DNA (mtDNA) play a central role in neuronal vulnerability and disease development (Borsche et al. 2021).

The mtDNA has unique characteristics, such as exclusive maternal inheritance, the absence of recombination, and a high mutation rate, which favor the accumulation of variants throughout life (Parakatselaki and Ladoukakis 2021a). Among these variants, single-nucleotide variants (SNVs) are particularly noteworthy, as they may be associated with impaired mitochondrial function. Previous studies have shown that SNVs in mitochondrial genes can affect oxidative phosphorylation, reactive oxygen species production, and apoptosis, processes intrinsically linked to neurodegenerative diseases (Sena-Dos-Santos et al. 2024; Kozin et al. 2025).

Despite the growing relevance of mtDNA, systematic research on the repertoire of mitochondrial SNVs (mtSNVs) in admixed populations remains scarce, particularly in Latin America, where haplogroups of indigenous ancestry predominate (de Souza et al. 2025). Furthermore, no studies have evaluated the functional impact of mtSNVs, the distribution of heteroplasmy, or the combined effects of multiple variants using polygenic risk scores (PRS) in PD in admixed populations(Rizig et al. 2023; Step et al. 2024; Saffie-Awad et al. 2025).

In this study, we characterized the repertoire of mtSNVs in peripheral blood samples from control individuals and people with PD from the Brazilian Amazon region. Our objective was to identify mitochondrial signatures that could serve as molecular markers of PD susceptibility. In addition, we proposed a mitochondrial polygenic risk score (mtPRS) weighted by the heteroplasmy levels of the risk variants.

## 2 Methods

### 2.1 Samples and study design

We carried out a cross-sectional observational study to investigate the repertoire of SNVs in the mtDNA of individuals with PD. A total of 192 participants were enrolled, including 109 patients diagnosed with PD (PD group) and 83 individuals without PD (CT group, controls), all recruited at the Movement Disorders Clinic of Hospital Ophir Loyola in Belém, Pará, Brazil. All participants underwent evaluation by the same movement disorders specialist (author B.L.S-L.), and both clinical and epidemiological data were collected. The PD and CT groups were matched for sex and age, with a maximum age difference of four years. Participants with acute or chronic infections, severe systemic illnesses, autoimmune disorders, or other neurological conditions were excluded.

All PD diagnoses were established according to the clinical criteria of the UK Parkinson’s Disease Society Brain Bank. Control participants were drawn from a voluntary cohort within the same population. The study protocol was approved by the Ethics Committee of Hospital Ophir Loyola (approval number 3.002.664).

### 2.2 Sample collection, DNA extraction, amplification, and sequencing

Peripheral blood was collected by venipuncture in EDTA tubes, stored at −20 °C, and used for DNA extraction. Extraction was carried out with the phenol-chloroform method, with modifications, and the KingFisher automated system. DNA concentration was measured with a Nanodrop 1000 (Thermo Fisher Scientific, Wilmington, DE, USA) and adjusted to 20 ng/µL.

The mitochondrial genome was amplified by PCR using 33 primer pairs (Cavalcante et al. 2019; de Souza et al. 2024). Libraries for mitochondrial genome sequencing were prepared using the Illumina Nextera XT DNA Library Preparation Kit in accordance with the manufacturer’s protocol. DNA integrity and fragment size distribution were assessed on the Agilent 2200 TapeStation using the D1000 High Sensitivity ScreenTape. Sequencing was carried out on Illumina MiSeq and NextSeq 500/550 platforms (Illumina Inc., Chicago, IL, USA). Ninety-six samples were sequenced on the MiSeq platform employing the MiSeq Reagent Kit v3 (600-cycle), while other 96 samples were sequenced on the NextSeq 500/550 using the High Output Reagent Kit v2.5 (75-cycle).

### 2.3 Bioinformatic analyses

#### 2.3.1 Sequencing data processing

Sequencing quality was evaluated with FastQC (v0.12.1) (Babraham Bioinformatics - FastQC A Quality Control tool for High Throughput Sequence Data) and MultiQC (v1.19) (Ewels et al. 2016). Low-quality bases, adapter sequences, and reads shorter than 36 nucleotides were removed using FastP (v0.23.4) (Chen et al. 2018). Processed .*fastq* files were aligned to the Revised Cambridge Reference Sequence (rCRS) with BWA (v0.7) (Li and Durbin 2009). Mapping, sorting, and duplicate removal were carried out with SAMTools (v1.15.1) (Danecek et al. 2021) and Picard (v2.27.5) (Broad Institute 2019). mtSNV detection and contamination analysis were performed with the mtDNA-Server pipeline (v2) (Weissensteiner et al. 2024) (Figure 1). Samples with contamination exceeding 10% were excluded, yielding 179 high-quality datasets (104 PD and 75 CT). All analysis scripts are available at GitHub (https://github.com/gilderlanio/Mitogenome-Variant-Calling) (Supplementary Figure 1).

**Figure 1.**
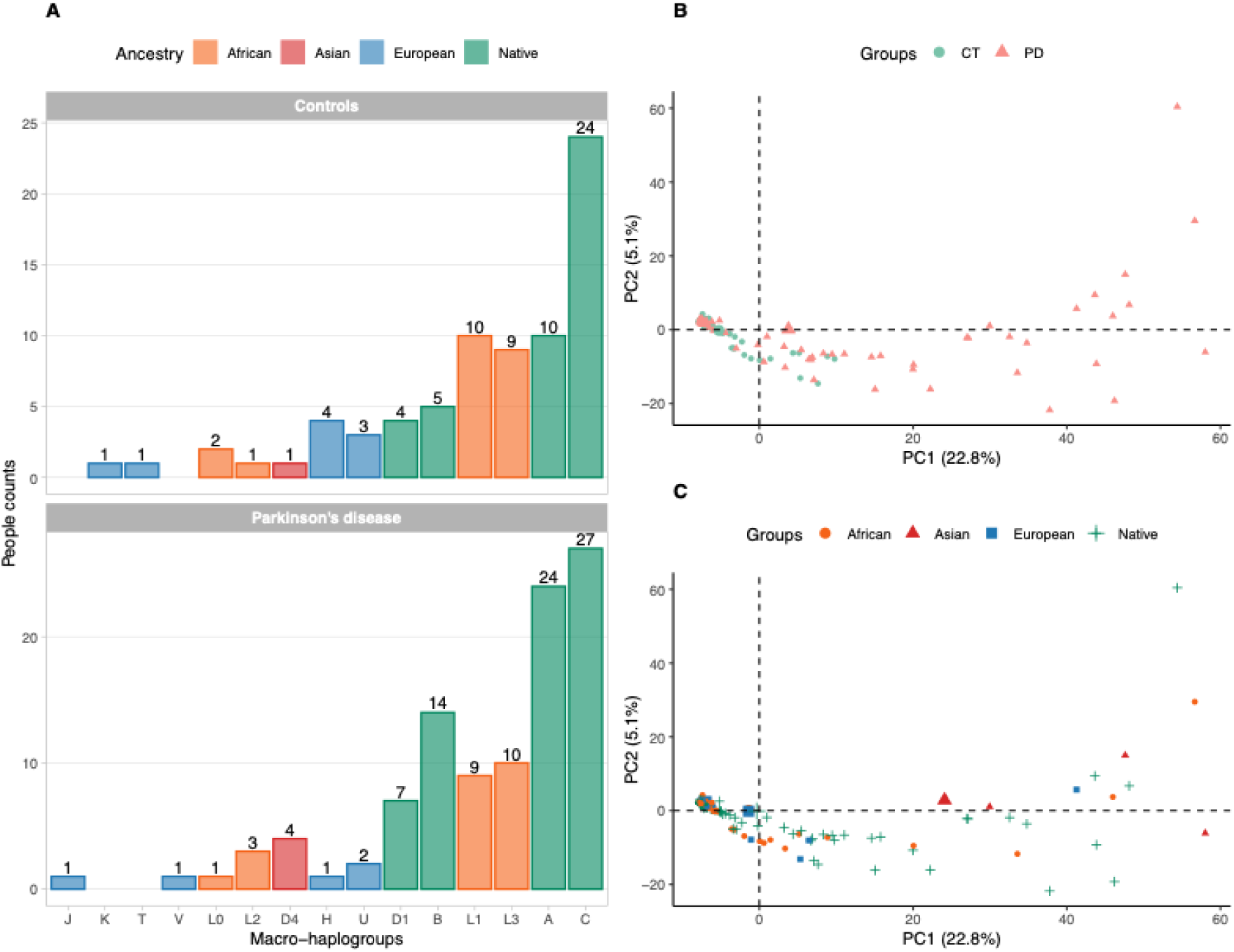
Flowchart of the bioinformatics pipeline for processing mtDNA sequencing data to identify and analyze mtSNVs. Steps include quality control, read alignment with the mitochondrial reference genome, variant identification, filtering, annotation, and downstream analysis.

#### 2.3.2 Quality and allele frequency filters

After excluding 13 individuals (5 PD and 8 CT) due to contamination levels above 10%, as determined by mtDNA-Server (v2), we applied a coverage filter, retaining only mtSNVs with a sequencing depth greater than 100x. Overall, sequencing yielded an average coverage of 1239.2x (Supplementary Figure 2). Next, we applied a heteroplasmy filter, selecting only heteroplasmic variants with frequencies above 0.05 (5%) and below 0.95 (95%).

After applying the coverage and heteroplasmy filters, we calculated the allelic frequency of the mtSNVs and set a minimum threshold of 0.01, retaining only recurrent heteroplasmic SNVs. Very rare variants were excluded.

#### 2.3.3 Statistical analysis

Comparisons of mtSNV burden, heteroplasmy levels across mitochondrial complexes, and mtPRS distributions CT and PD groups were performed using the Wilcoxon rank-sum test. Principal component analysis (PCA) was conducted using a binary absence/presence matrix of unique mtSNVs to capture population structure.

Logistic regression was used to identify mtSNVs associated with PD, adjusting for age, sex, population structure, and sequencing batch (Seq-batch). Collinearity among covariates was evaluated using the Variance Inflation Factor (VIF), a widely used metric to quantify multicollinearity in regression models (O’brien 2007). Based on this assessment, only the first principal component (PC1) was retained in the final model to ensure statistical robustness and model stability.

For each mtSNV, odds ratios (ORs) and 95% confidence intervals (CIs) were estimated. OR > 1 indicated increased risk, whereas OR < 1 indicated a protective effect. Multiple testing correction was applied using the Benjamini–Hochberg procedure, and associations with a false discovery rate (FDR) < 0.05 were considered statistically significant.

##### 2.3.3.1 Mitochondrial Polygenic Risk Score

The PRS is a metric computed as a weighted sum of individual genotypes, with weights assigned to each variant. It is an additive linear model, in which each variant contributes independently and cumulatively to the total risk. Thus, the PRS does not involve derivation or complex modeling in its formulation and is essentially a simple linear combination of genetic effects (Lewis and Vassos 2020).

Therefore, it is a metric that summarizes the cumulative effect of multiple genetic variants on disease risk. Traditionally, the PRS is applied to biallelic nuclear variants, in which each individual carries 0, 1, or 2 copies of the risk allele. In this approach, the PRS is defined as:

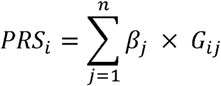

Where:

- *b_j_* corresponds to the estimated effect of variant *j*, representing the logarithm of the odds ratio (log-odds) associated with the presence of the risk allele.
- *G_ij_* is the individual’s genetic load for variant *j*, assuming values 0, 1 or 2 depending on the number of copies of the risk allele present.
- *n* is the total number of variants included in the score.

Unlike the nuclear genome, mitochondria can host multiple copies of both wild-type and mutated mtDNA. This condition, called heteroplasmy, refers to the relative proportion of mtDNA molecules that carry a specific variant. (Parakatselaki and Ladoukakis 2021b). Thus, instead of considering a fixed genotype (0, 1, or 2 copies), we proposed using the heteroplasmy rate as a continuous measure of allelic load. Therefore, the PRS calculation was extended for mtDNA (mtPRS) as follows:

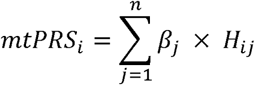

Where:

- *b_j_* represents the effect of variant *j* on disease risk.
- *H_ij_* corresponds to the heteroplasmy rate of variant *j*, ranging from 0 (no mutated mitochondrial copies) to 1 (all mutated copies, or homoplasmy);
- *n* is the number of mitochondrial variants considered in the score.

Multiplying *b_j_* by heteroplasmy (*H_ij_*) reflects the true intensity of each variant’s contribution. Risk variants present in high heteroplasmy have a proportionally greater impact on the PRS, while variants in low heteroplasmy have a reduced effect. Similarly, variants with a protective effect (negative *b_j_* values) attenuate the score in proportion to their relative frequency in the individual’s mtDNA.

Thus, an mtPRS was calculated for each individual. We then performed a logistic regression, adjusted for sex and age, with mtPRS as the independent predictor to assess the association between each mtPRS and PD. The model’s discriminatory ability was evaluated using ROC curve analysis. Predicted probabilities from a logistic regression model were used to construct the ROC curve and estimate AUC. To assess the robustness of predictive performance and reduce optimistic bias, 5-fold cross-validation was performed, in which the model was trained on independent subsets of the data and evaluated on the remaining subsets. The AUC was estimated as the mean AUC across the 5-fold validation sets. All statistical analyses were performed using RStudio.

#### 2.3.4 Functional annotation and pathogenicity prediction

Additionally, we used VEP and Apogee 2 for functional annotation and pathogenicity prediction, respectively. We chose Apogee because it performs better for mitochondrial variants (Bianco et al. 2023). Furthermore, we used MITOMASTER (July 15, 2025) to assess conservation levels (Brandon et al. 2009).

## 3 Results

### 3.1 Demographical and clinical characteristics

A significant difference in sex distribution was observed between the PD and CT groups (p = 0.023), with males comprising a larger proportion of the PD group (67.3%) compared to controls (49.3%). Family history was also significantly associated with PD, reported by 24.0% of patients and 5.3% of controls. The mean age did not differ significantly between groups (*p* = 0.419). Among individuals with PD, tremor was the most frequent initial motor manifestation (63.5%), and the tremor-dominant phenotype was the most common (42.3%). Most patients were in the early stages of the disease (Hoehn & Yahr stages 1–2, 76.0%). The median duration of levodopa treatment was 5 years, with a median daily dose of 662.5 mg (Supplementary Table 1).

### 3.2 Mitochondrial ancestry distribution indicates population stratification

Mitochondrial ancestry analysis revealed a predominance of macrohaplogroups of Native American origin (A, B, C, and D1) among the individuals evaluated, both in PD patients and controls, followed by haplogroups of African (L0, L1, L2, and L3), Asian (D4), and European (J, K, T, V, H, and U) origin, reflecting the characteristic genetic profile of the Amazonian population studied (Figure 1A). PCA, based on the presence/absence of mtSNVs, revealed broad overlap between the case and control groups, without distinct clustering, indicating that mitochondrial variability does not differentiate individuals by clinical condition (Figure 1B). The first two principal components accounted for 22.8% (PC1) and 5.1% (PC2) of the total variance. When individuals were classified by ancestry, greater dispersion was observed among those with Native American, African, and European components, reflecting the high degree of admixture in the Brazilian Amazonian population (Figure 1C).

### 3.3 Increased incidence of low-heteroplasmy mtSNVs in Complex I, IV, V, and rRNA genes in PD

Across individuals, we identified 12,481 SNVs (PD: 9,793; CT: 2,688), with a predominance of variants with heteroplasmy levels below 0.50 (50%), located mainly in ribosomal RNA (rRNA) genes (*p* = 1.4 × 10^-5^). Among protein-coding genes, the burden was most evident in Complex I genes (*p* = 0.0056) (mainly *ND3*, *ND4*, *ND5*, and *ND6*), followed by Complex IV genes (*p* = 2.1 x 10^-6^) (*COX2* and *COX3*). The burden was more evident in individuals with PD (Figure 2 and Supplementary Figure 3).

**Figure 2.**
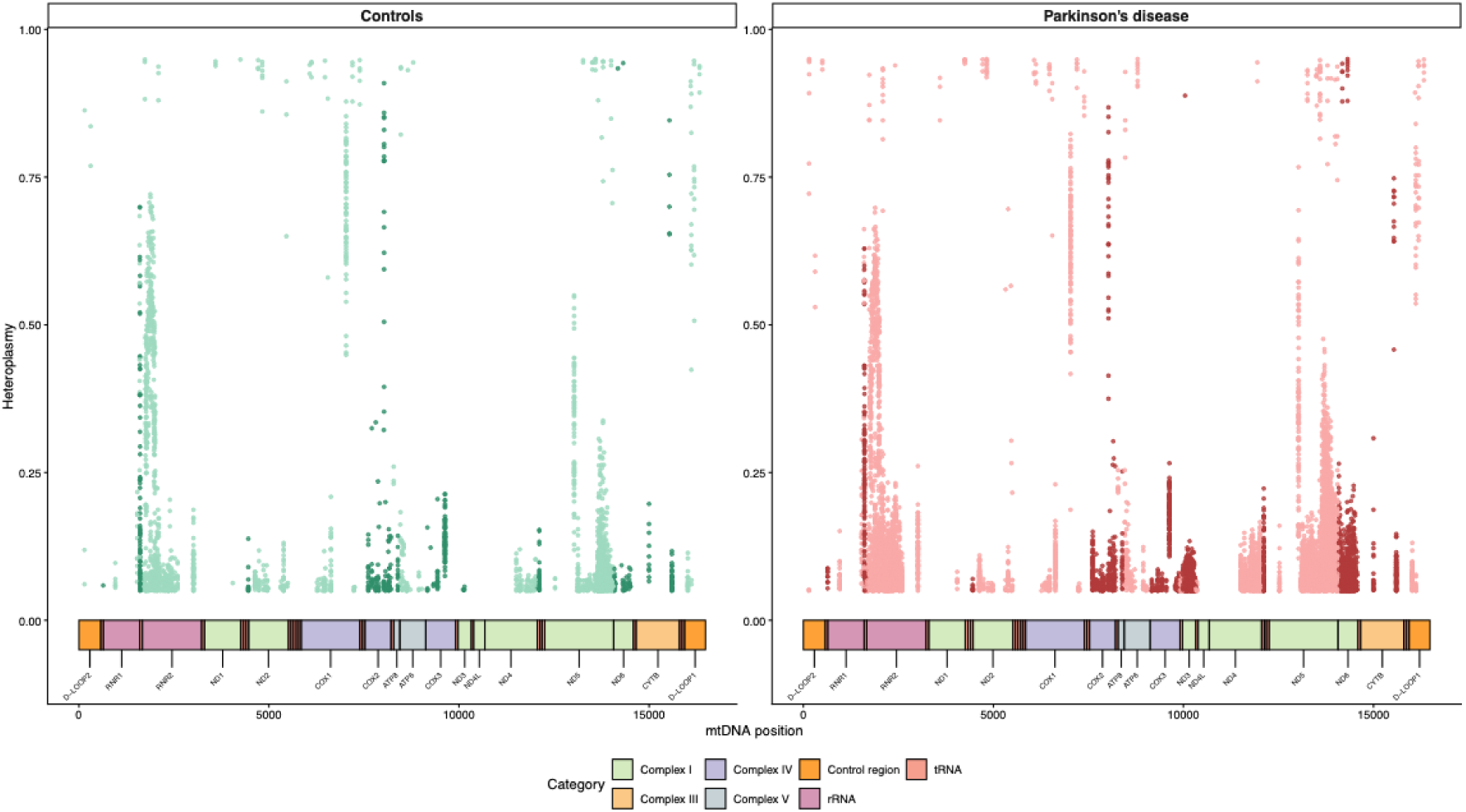
Distribution of mitochondrial macrohaplogroups by ancestry and principal component analysis (PCA) of people with Parkinson’s disease (PD) and controls (CT). **(A)** Distribution of mitochondrial macrohaplogroups by ancestry among people with PD and CT, showing a predominance of lineages of Native American origin. **(B)** PCA based on the presence/absence of mtSNVs, classifying individuals according to the PD and CT groups, showing broad overlap between groups and the absence of distinct clusters. **(C)** PCA based on the presence/absence of mtSNVs, classifying individuals according to ancestry, highlighting greater dispersion among those with Native American, European, and African components.

### 3.4 Low-heteroplasmy mtSNVs in *RNR2*, *COX1*, *COX2*, and *ND4* contribute to increased mtPRS in PD

We identified 536 unique mtSNVs, of which 321 were shared between individuals with PD and CT, 214 were exclusive to PD, and one was exclusive to CT (Figure 3A). Association analyzes were conducted using logistic regression, and model selection was based on VIF assessment. In the full model including Age, Sex, PC1, PC2, PC3, and sequencing batch, high VIF values were observed for PC1 (VIF = 4.417) and PC2 (VIF = 5.188), indicating strong collinearity. An intermediate model including Age, Sex, PC1, PC2, and sequencing batch showed increased multicollinearity, with VIFs of 5,810 for PC1 and 6,724 for PC2. In contrast, the reduced model including Age, Sex, PC1, and sequencing batch presented VIF values close to 1 (Age = 1.062, Sex = 1.028, PC1 = 1.737, and sequencing batch = 1.795), indicating minimal collinearity and greater statistical robustness. Therefore, only PC1 was retained as a population structure covariate in the final model (Supplementary Table 2).

**Figure 3.**
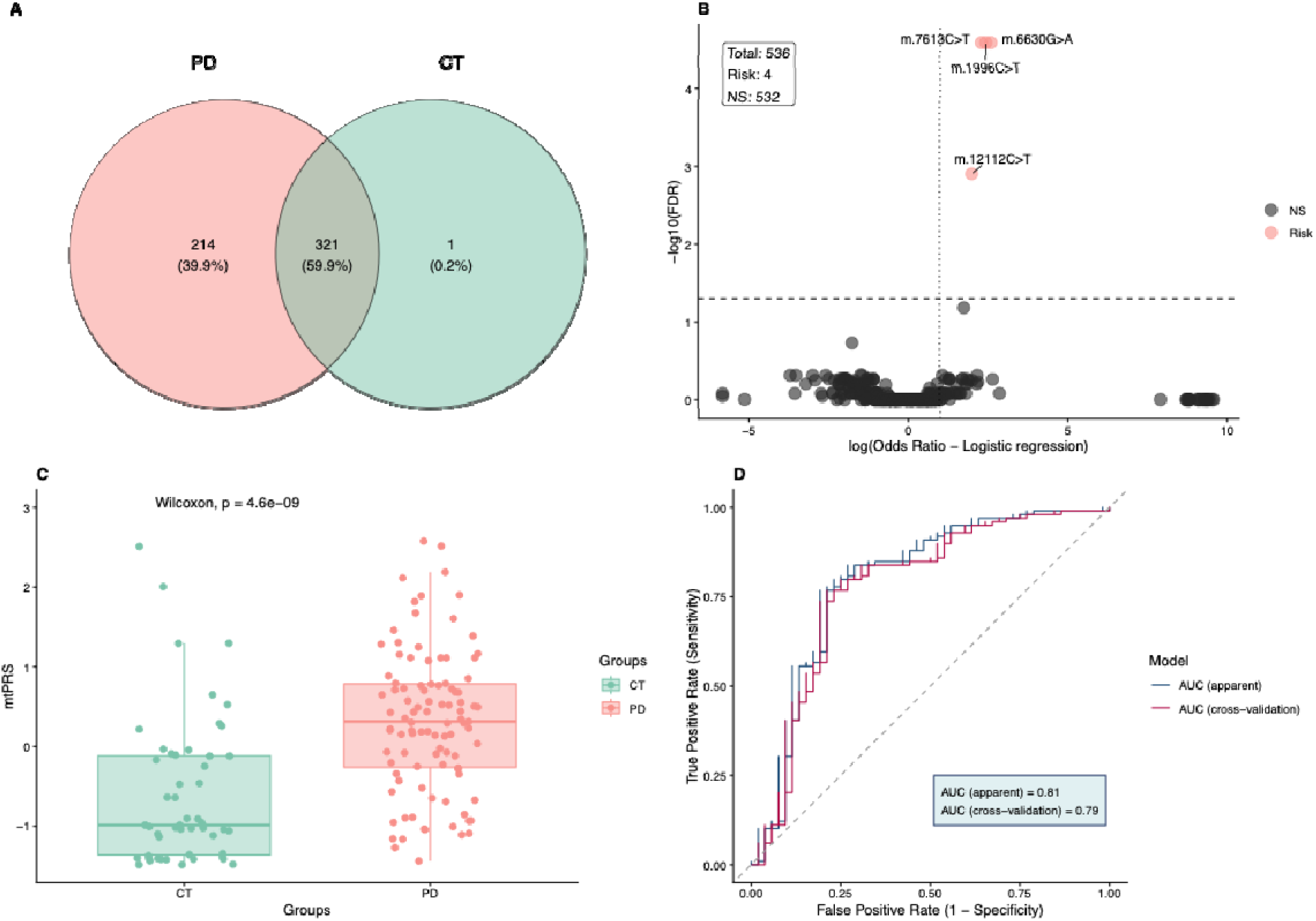
Distribution of heteroplasmy levels for mitochondrial mtSNVs identified in individuals with PD and CT groups across the mtDNA.

Using this final model, four mtSNVs were significantly associated with PD (m.1996C>T, m.6630G>A, m.7613C>T, and m.12112C>T; FDR < 0.05; OR > 1), with exact odds ratios and 95% confidence intervals (CI) reported in Supplementary Table 3 (Figure 3B). Using the mtSNVs at risk, we calculated the mtPRS and observed a statistically significant difference between the groups (*p* = 4.6 × 10_^9^), with higher scores in patients with PD (Figure 3C). The analysis of the mtPRS constructed from risk mtSNVs showed a significant association with PD (OR = 3.64; 95% CI: 2.28 - 6.22; *p* = 3.76 × 10_^7^; *FDR* = 1.12 × 10_^6^). Furthermore, mtPRS demonstrated a significant association with PD (OR = 3.64; 95% CI: 2.28 - 6.22; *p* = 3.76 × 10__; *FDR* = 1.12 × 10__). The model demonstrated good discriminative performance, with an AUC of 0.81 (95% CI: 0.72 - 0.88), indicating a strong ability to distinguish between cases and controls. To assess the robustness of the model and reduce optimistic bias, a 5-folds cross-validation was performed, resulting in an AUC of 0.79 (95% CI: 0.73 - 0.92), a value very close to that observed in the apparent analysis (Figure 3D).

### 3.5 Functional and evolutionary features of PD-associated mtDNA SNVs

The m.1996C>T variant, located in the *RNR2* gene (rRNA), showed a heteroplasmy distribution of 0.113 (IQR: 0.079-0.139) in people with PD and 0.096 (IQR: 0.067-0.123) in CT, with no statistically significant difference (p = 0.072). This variant exhibited 86.67% conservation relative to the reference base and is not cataloged in any databases. The missense mtSNV m.6630G>A in the *COX1* gene (Complex IV) showed a higher heteroplasmy level in the PD group (0.096; IQR: 0.076–0.117) than in CT (0.059; IQR: 0.052–0.105; p = 0.00004). This position is 100% conserved and was described as pathogenic by Apogee (score = 0.68) (Supplementary Table 4).

The mtSNV m.7613C>T in the *COX2* gene, classified as a stop-gain variant, showed heteroplasmy rates of 0.082 (IQR: 0.071–0.102) in PD and 0.075 (IQR: 0.057–0.090) in CT, with no statistically significant difference (p = 0.332). It is a variant that occurs at a position of high evolutionary conservation (97.78%) and is undescribed. Finally, the synonymous SNV rs28695839 (m.12112C>T) in the *ND4* gene (Complex I) showed higher heteroplasmy rates in DP (0.091; IQR: 0.069–0.132) than in controls (0.068; IQR: 0.058–0.078; p = 0.00006) and 97.78% of conservation (Supplementary Table 4).

## 4 Discussion

This study identified mitochondrial molecular signatures in peripheral blood samples from individuals with PD originating from a highly admixed population in the Amazon region of northern Brazil. Due to the intense historical process of admixture, this region presents multiple ancestral contributions. However, a predominance of Native American ancestry is observed (Kehdy et al. 2015; Nunes et al. 2025), as evidenced by the high frequency of mitochondrial haplogroups A, B, C, and D, as shown by our results and by Souza et al. (de Souza et al. 2025). The investigated population has been historically underrepresented in genomic studies, particularly in the context of neurodegenerative diseases. Thus, this work aims to reduce these knowledge gaps and provide new insights into Parkinson’s disease among admixed individuals with substantial Native American ancestry, thereby broadening understanding of the genetic diversity associated with the disease.

In the mtDNA of these individuals, we observed a higher burden of mtSNVs in Complex I, IV, and V genes, as well as in rRNA genes, in people with PD compared with controls. The increased mtSNV burden in PD may reflect a cycle involving chronic inflammation, excessive pro-inflammatory cytokine release, increased oxidative stress, and heightened susceptibility to mtDNA damage. This process likely contributes to impaired mitophagy, which in turn compromises inflammation resolution and promotes its persistence (Sa et al. 2022; Orekhov et al. 2025). This mechanistic hypothesis is based on the premise that PD is a systemic inflammatory condition that extends beyond its well-recognized neuroinflammatory focus the substantia nigra (Joshi and Singh 2018). Accordingly, it is well established that there is an increased concentration of pro-inflammatory cytokines (IL-1β, TNF-α, and IL-6) and activation of inflammatory pathways in the peripheral blood of individuals with PD (Qu et al. 2023), which may increase oxidative stress levels (Yang et al. 2007; Kim et al. 2010). Previous studies have reported oxidative damage to mtDNA in PD (Migliore et al. 2002). In addition, pro-inflammatory cytokines can stimulate NADPH oxidase assembly, leading to a marked increase in reactive oxygen species (ROS) production in peripheral blood mononuclear cells, a phenomenon previously described in PD (Liu et al. 2022). IL-17A secretion by Th17 cells, a predominantly pro-inflammatory mediator, has been associated with enhanced neurodegeneration and with abnormal ROS accumulation (Chen et al. 2020).

It is important to note that the higher proportion of mtSNVs observed in people with PD compared to controls should be interpreted with caution. The mtDNA recovered from peripheral blood samples originates predominantly from peripheral blood mononuclear cells and platelets (Picard 2021). As quantitative data on potential differences in the relative abundance of these cell populations between groups were not available, variations in cellular composition may have led to an over- or underestimation of the number of mtSNVs detected.

Additionally, within the mtSNV burden identified in Complex IV, we detected two mtSNVs: a *stop-gained* variant in *COX2* and a pathogenic *missense* variant in *COX1*. These variants may lead to inactivation of Complex IV. Silencing of this complex renders macrophages more pro-inflammatory, as inhibition of *COX* genes in macrophages has been shown to increase IL-1β, IL-6, and TNF-α expression, promote polarization toward the M1 phenotype, and enhance phagocytic activity. (Angireddy et al. 2019), as already described in the DP (Moehle and West 2015). In Complex I, a synonymous mtSNV in *ND4* was associated with PD risk. Synonymous mtDNA variants may exert functional effects primarily by altering translational efficiency and kinetics, despite not changing the aminoacid sequence. In mitochondria, the limited tRNA pool and codon bias amplify these effects, as codon changes can slow ribosomal translation, disrupt co-translational protein folding, and impair the assembly or stability of respiratory chain complexes (Lareau et al. 2025), including Complex I.

In *RNR2*, mtSNVs may affect the structure of the mitochondrial 16S rRNA, thereby altering mitoribosome assembly or stability. Previous studies have shown that variants in mitochondrial rRNAs can reduce mitochondrial translation rates and increase mitochondrial stress, even at moderate or low levels of heteroplasmy (Greber and Ban 2016; Ferrari et al. 2021). In addition, *RNR2* exhibits high population variability and is frequently enriched in mtSNVs due to the lack of direct selective pressure on aminoacid sequences, in contrast to protein-coding genes. This characteristic renders the gene particularly sensitive to contextual effects, in which variants that appear neutral may become deleterious when combined with other genetic factors, environmental exposures, or aging-related processes (Wallace 2015). Another relevant hypothesis is that mtSNVs in *RNR2* may act as risk-modulating variants rather than as direct causal variants. In this scenario, they would contribute to a dysfunctional mitochondrial environment that, together with mtSNVs in protein-coding genes, increases overall mitochondrial vulnerability, as previously described in complex diseases (Ivanova et al. 2021; Bibi et al. 2023).

In addition, the variants associated with PD predominantly occur at low levels of heteroplasmy, likely below the biochemical threshold required to produce detectable pathogenic effects when considered individually. However, when combined with other mtDNA variants, these alterations may act in a cumulative manner and contribute to the manifestation of PD (Smith et al. 2024). In this context, by constructing mtPRS from risk-associated mtSNVs, we observed significantly higher scores in individuals with PD, with a strong statistical association and good discriminatory performance.

Nevertheless, it is important to note that the proposed mtPRS was built using novel mitochondrial variants identified in a population with predominant Native American ancestry, which precluded direct validation of its performance in independent cohorts from other populations. Despite this limitation, the adopted approach represents a relevant methodological advance, as mtSNVs remain largely overlooked in PRS calculations, which have focused primarily on nuclear variants (Schwarzerova et al. 2024). In this regard, the mtPRS presented here stands out by explicitly incorporating mitochondrial genetic variability as a central component of risk prediction.

The relevance of this approach becomes even more evident considering previous evidence showing that PRS based on nuclear variants associated with PD, largely derived from European populations, exhibit substantial loss of performance when applied to Latin American cohorts (Loesch et al. 2022). Similarly, a PRS constructed from variants in nuclear genes involved in mitochondrial pathways demonstrated significant association and good predictive performance in European and Ashkenazi populations, but not in African or Asian populations (Ooi et al. 2025). These findings underscore the population-dependent modularity of the genetic architecture of PD.

Within this context, the mtPRS proposed in this study is specifically tailored to mtDNA variants and demonstrates consistent predictive performance in an underrepresented population with predominantly Native American ancestry. Collectively, these results emphasize the importance of incorporating population diversity and mitochondrial genetic variation into future genetic risk models for PD, contributing to the development of more equitable and biologically informed predictive tools.

In summary, this study provides evidence that novel low-heteroplasmy mtSNVs, particularly in *RNR2*, *COX1*, *COX2*, and *ND4*, may contribute additively to the risk of PD in admixed populations. However, some limitations should be considered: the analyses were performed on mtDNA derived from peripheral blood, which may not fully reflect mitochondrial processes in tissues directly affected by the disease, such as the brain. Furthermore, the observational nature of the study prevents direct causal inferences. Future studies that integrate functional approaches, analyses of specific tissues, and independent cohorts of different ancestries will be essential to validate these findings and clarify the mechanisms by which low-heteroplasmy mitochondrial variants modulate susceptibility to PD.

## Data Availability Statement

Mitochondrial genome sequencing data were deposited in the European Nucleotide Archive (ENA) under accession number PRJEB74357, and the raw data are also available through the mtDNA Network platform (https://apps.lghm.ufpa.br/mtdna/).

## Data Availability

Mitochondrial genome sequencing data were deposited in the European Nucleotide Archive (ENA) under accession number PRJEB74357.

## Acknowledgments

We sincerely thank all individuals who participated in this study and acknowledge the financial support provided by CNPq, CAPES, PROPESP/UFPA, and FAPESPA. We also highlight the relevance of exploring genetic mechanisms in diverse and historically underrepresented populations. We hope that the results of this research will contribute to advances in public health.

## Author Contribution Statement

GBM was responsible for the methodology, formal analyses, data curation, data visualization, and preparation of the original manuscript. FGS, CSS, TPS, GCC, and AVSF contributed to the methodological aspects of the study. MEA and MVBS participated in the revision and editing of the manuscript. LCCM contributed to data visualization and software support. BLSL provided resources, contributed to manuscript revision, and supervised the study. ÂRS was responsible for funding acquisition, project management, and supervision. GSA led the conceptualization of the study and contributed to formal analyses, manuscript writing and revision, project administration, and supervision. All authors reviewed and approved the final version of the manuscript.

## Ethical Approval

The study was approved by the Hospital Ophir Loyola Ethics Committee (number 3.002.664).

**SNV distribution, group associations, and mitochondrial polygenic risk score (mtPRS)**. **(A)** Distribution of the remaining 536 SNVs after the MAF > 0.01 filter between CT and DP. **(B)** Volcano plot representing the results (adjusted p-value: FDR; Odds ratio) of the logistic regression adjusted for sex, age, PC1, and sequencing batch in the 536 unique SNVs. **(C)** Comparison of mtPRS distribution between people with PD and control individuals. **(D)** ROC curve of the logistic regression model fitted for mtPRS, age, and sex, evaluating the discrimination between cases and controls. The apparent curve represents the performance of the model fitted to the complete dataset (train and test sets), whereas the cross-validation curve reflects the average performance on data not used in fitting.

