## Supplementary Material for "Novel mitochondrial variants associated with Parkinson’s disease reveal a high heteroplasmy-weighted polygenic risk score in Brazilians"

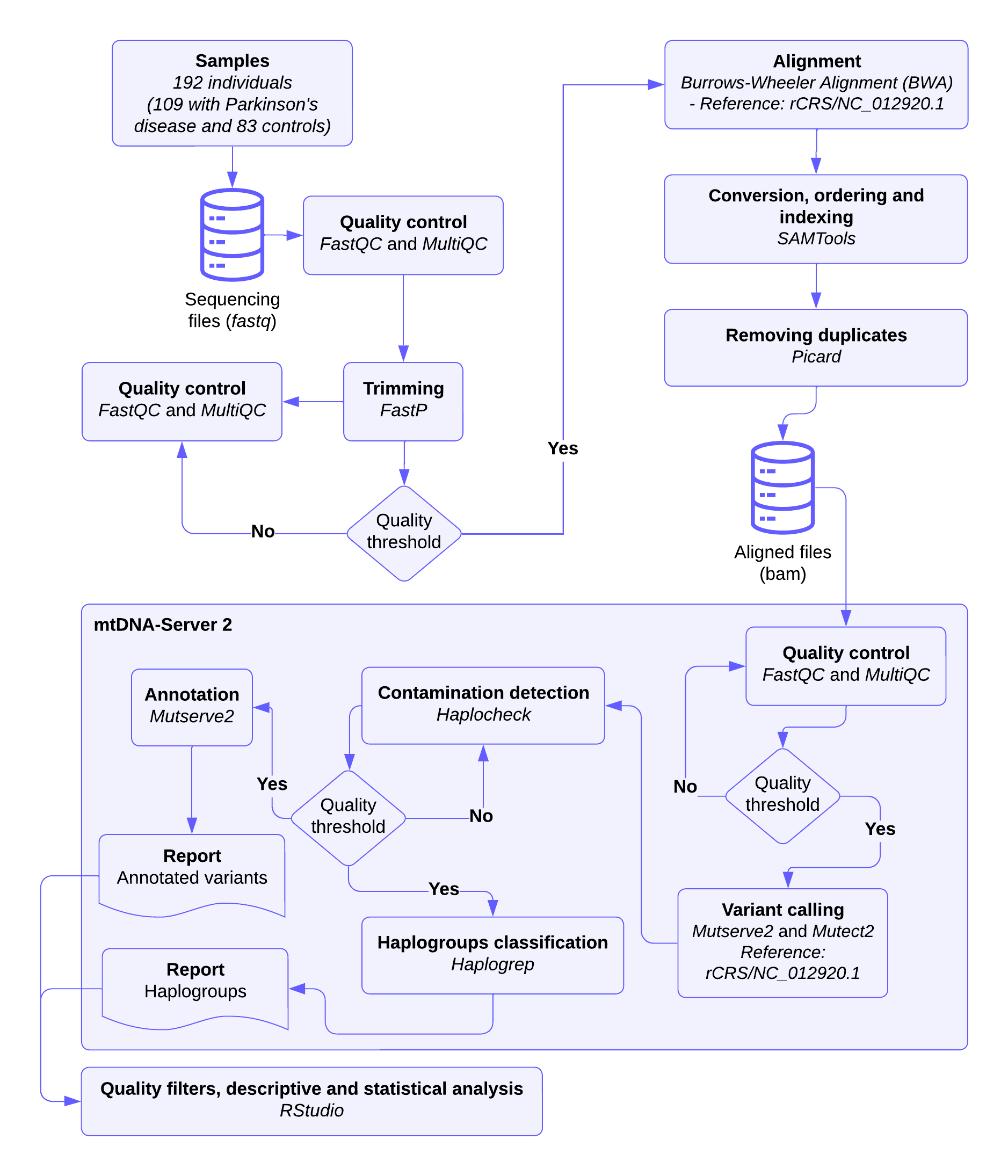

**Supplementary Figure 1. Flowchart of the bioinformatics pipeline for processing mtDNA sequencing data to identify and analyze mtSNVs.** Steps include quality control, read alignment with the mitochondrial reference genome, variant identification, filtering, annotation, and downstream analysis.

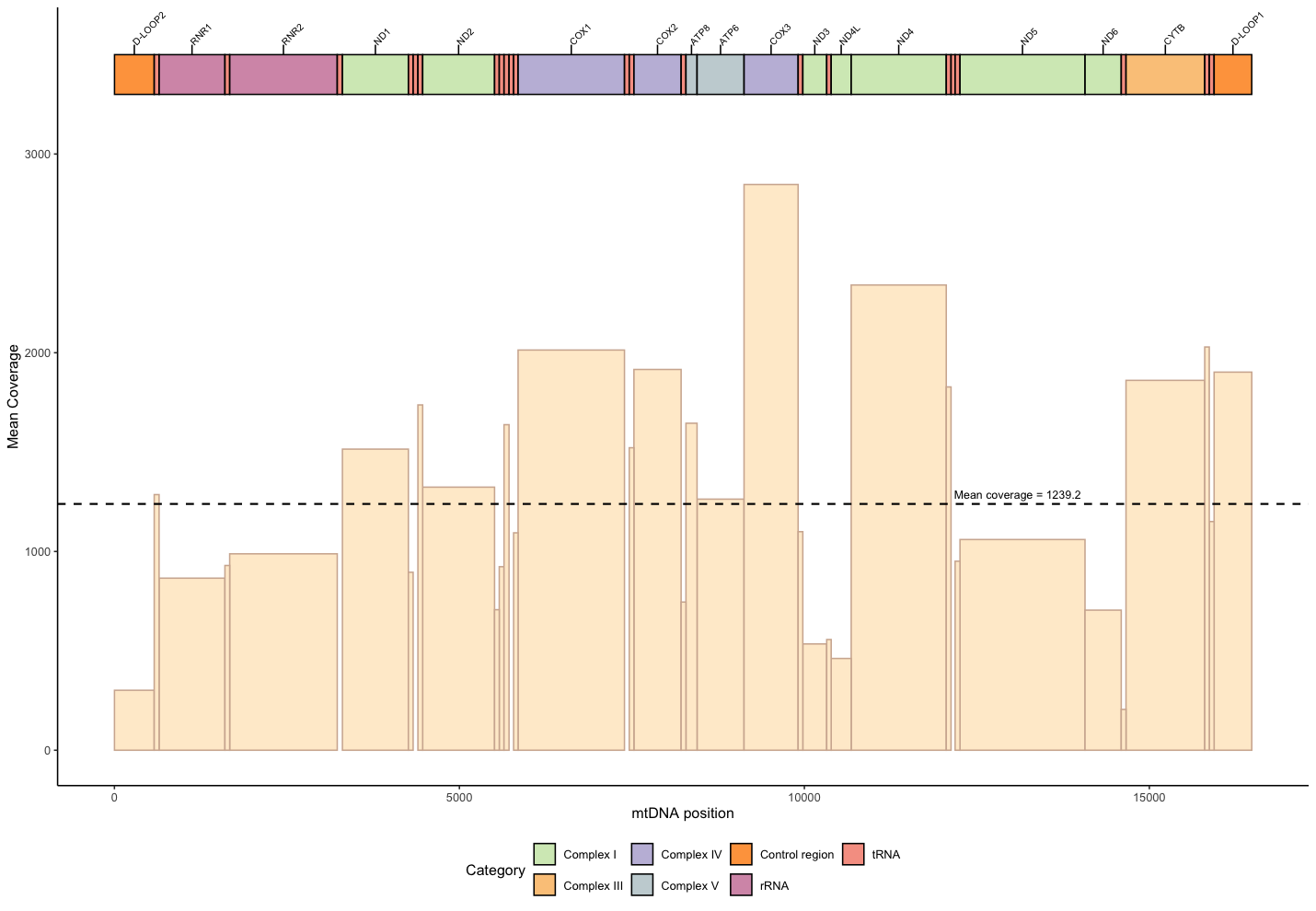

**Supplementary figure 2.** Distribution of sequencing coverage depth along mtDNA.

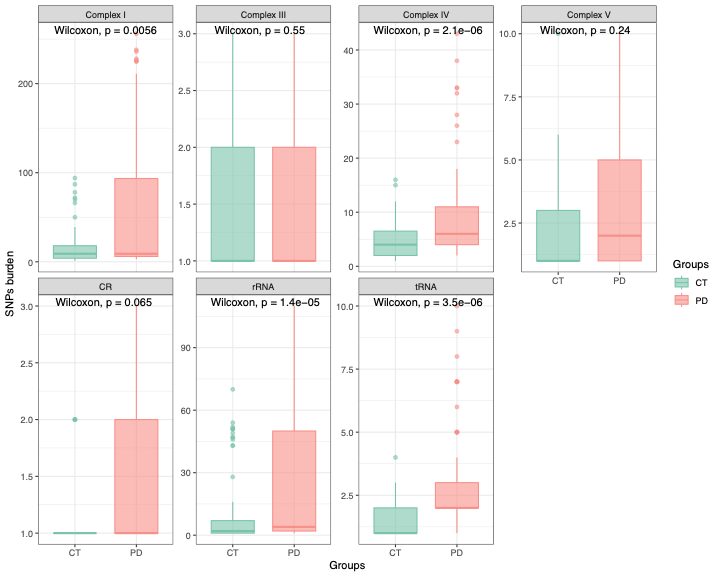

**Supplementary figure 3.** Statistical comparison of SNV burden per mitochondrial complex between people with Parkinson's disease and controls.
